# The relationship between anxiety and levels of Alzheimer’s disease plasma biomarkers

**DOI:** 10.1101/2024.07.09.24310168

**Authors:** Mark A. Bernard, Allal Boutajangout, Ludovic Debure, Wajiha Ahmed, Anthony Q. Briggs, Carolina Boza-Calvo, Alok Vedvyas, Karyn Marsh, Omonigho M. Bubu, Ricardo S. Osorio, Thomas Wisniewski, Arjun V. Masurkar

## Abstract

Anxiety is highly prevalent in Alzheimer’s disease (AD), correlating with CSF/PET biomarkers and disease progression. Relationships to plasma biomarkers are unclear. Herein, we compare levels of plasma biomarkers in research participants with and without anxiety at cognitively normal, mild cognitive impairment, and AD dementia stages. We observed significantly higher plasma tau/Aβ42 ratio in AD participants with anxiety versus those without, but did not observe differences at other stages or plasma biomarkers. No such relationships were evident with depression. These results support a unique pathophysiological relationship between anxiety and AD that can be reflected in plasma biomarkers, suggestive of heightened neurodegeneration.

## INTRODUCTION

Neuropsychiatric symptoms (NPS) are common in Alzheimer’s disease (AD) and AD-related dementias (ADRD) [1, 2]. In particular, anxiety is highly prevalent in individuals with AD even at early stages, varying between 10-49% among those with subjective cognitive decline, and 10-70% in individuals with mild cognitive impairment (MCI), and is associated with faster disease progression [3].

This relationship between anxiety and AD is also seen at the biomarker level, including structural magnetic resonance (MRI), F-fluorodeoxyglucose positron emission tomography (FDG-PET), and amyloid/tau measured via cerebrospinal fluid and PET [3]. However, the correlation of anxiety to plasma AD biomarkers is unclear. Plasma biomarkers confer several potential advantages for evaluating AD pathogenesis. There is evidence that changes in plasma biomarkers may preview changes in CSF or PET biomarkers, thus potentially enabling earlier insight into the relationship of anxiety with disease progression at early stages. In addition, assessment of proteinopathy in the periphery can offer a more in-depth investigation into dynamics leading to amyloid/tau deposition in the brain [4, 5]. From a practical perspective, if anxiety is to be utilized as a prescreen for further biomarker investigation in vulnerable populations, plasma biomarkers offer a more cost-effective and less invasive means to assess AD risk at a biological level.

In this study, we performed a cross-sectional analysis on a research cohort of older adults to test the hypothesis that plasma AD biomarker levels differ between anxious and non-anxious participants across the AD spectrum. We leveraged SIMOA technology to acquire high-fidelity measurements of plasma Aβ42, Aβ40, NfL, and total tau. Additionally, we assessed whether potential differences in plasma AD biomarker relationships were unique to anxiety by also investigating their correlations to depression.

## MATERIALS AND METHODS

### Study participants

A cross-sectional analysis was performed on a deidentified dataset of participants evaluated at the NYU Alzheimer’s Disease Research Center (ADRC) between 2006 and 2019. The NYU ADRC study is approved by the New York University Grossman School of Medicine Institutional Review Board. Participants were community-dwelling older adults living in the greater New York City metropolitan area, recruited through community events and physician- or self-referrals. NYU ADRC inclusion criteria were age > 50, a study partner, fluency of participant and study partner in English or Spanish, and willingness to undergo phlebotomy and MRI. Exclusion criteria included significant non-AD/ADRD neurological disease, HIV/AIDS, organ transplantation, organ failure, significant autoimmune disease, significant psychiatric disease, malignancy within the last 5 years, and alcohol/drug abuse. The present study includes participants whose study partners gave responses to questions for anxiety and depression on the NPI-Q, and who had data for at least one AD plasma biomarker measured (n = 197).

### Clinical measurements

Participants underwent clinical interviews, physical examinations, and were administered psychometric testing and other instruments according to the National Alzheimer’s Coordinating Center (NACC) Uniform Data Set, in addition to NYU-specific assessments. These data were synthesized into a consensus diagnosis by ADRC Clinical Core clinicians to determine AD stage (e.g. cognitively normal, MCI, mild AD dementia). Neuropsychiatric symptoms were assessed by study partner report via the Neuropsychiatric Inventory Questionnaire (NPI-Q) for anxiety, depression, and other symptoms [6]. Participants were considered anxious or depressed if study partners answered affirmatively to the respective NPI-Q items. Blood was also collected and stored for evaluation of APOE genotype and measurement of AD plasma biomarkers.

### Measurement of blood plasma biomarkers

Blood samples were collected in 10 mL EDTA tubes, gently inverted, and centrifuged at 3000 rpm for 15 minutes at room temperature. The plasma was extracted into propylene low-binding tubes, which were promptly stored at -80° C. Plasma samples were centrifuged 10.000 g for 5 minutes before use. Plasma biomarker levels of Aβ42, Aβ40, total tau, and NfL were measured using a Quanterix Single Molecule Array (SIMOA) HD-1 Analyzer and related kits. Assays were performed in duplicate using two internal standards and calibrated with an 8-point calibration curve.

### Statistical Analysis

Statistical analysis was performed with GraphPad Prism. Chi-squared tests were used to compare categorical and demographic data between groups. Unpaired t-tests were performed to compare means of continuous variables (measured plasma biomarker levels) between anxious and non-anxious groups of participants. Similar analyses were conducted in participants with depression.

## RESULTS

Demographic and genetic data for the 197 participants are summarized in Table 1. Of 197 participants, 103 were cognitively normal (CN), 53 were at mild cognitive impairment (MCI), and 41 were at the AD stage. Anxiety had a prevalence of 14%, 48%, and 24% at CN, MCI, and AD stages, respectively among this cohort. There were no statistically significant differences among anxious and non-anxious groups within a particular stage, nor between stages among anxious/non-anxious groups (Table 2).

**Table 1.** Demographic and psychometric characteristics of the study population.

| Stage | CN |  | MCI |  | AD |  |
| --- | --- | --- | --- | --- | --- | --- |
|  | Anxious<br>(n=16) | Non-Anxious<br>(n=87) | Anxious<br>(n=25) | Non-Anxious<br>(n=28) | Anxious<br>(n=10) | Non-Anxious<br>(n=31) |
| **Demographic** |  |  |  |  |  |  |
| **Age, years (SD)** | 67.94<br>(5.38) | 72.26<br>(7.81) | 75.91<br>(10.18) | 77.53<br>(8.98) | 78.52<br>(10.95) | 82.94<br>(9.61) |
| **% Women** | 75.00 | 65.52 | 72.00 | 85.71 | 80.00 | 58.06 |
| **%Non-Hispanic**<br>**White** | 87.50 | 87.36 | 92.00 | 78.57 | 40.00 | 83.87 |
| **Education, years**<br>**(SD)** | 14.88<br>(4.15) | 17.17<br>(2.36) | 16.84<br>(2.34) | 16.07<br>(3.39) | 12.90<br>(5.71) | 12.83<br>(5.24) |
| **% ApoE4 carrier** | 25.00 | 26.44 | 40.00 | 25.00 | 50.00 | 25.81 |

**Table 2.** Comparisons of psychometric instrument scores between anxious and non-anxious AD participants (values reported as test scores +/- SD).

| Test/Group | AD-anxiety | AD+anxiety | p-value |
| --- | --- | --- | --- |
| **MMSE** | 20.77 (5.56) | 21.22 (3.19) | 0.8190 |
| **Animals** | 11.90 (3.85) | 10.78 (4.52) | 0.4687 |
| **Vegetables** | 7.79 (2.92) | 7.33 (2.12) | 0.6652 |
| **Trails A** | 72.19 (39.75) | 76.63 (42.41) | 0.7769 |

We compared the levels of each plasma biomarker (Aβ40, Aβ42, NfL, tau) and relevant ratios (Aβ42/40, tau/Aβ42) between anxious and non-anxious participants across each AD stage. The tau/Aβ42 ratio was the only measure to show statistically significant findings (Figure 1A), with plasma tau/Aβ42 ratios elevated by 66% between anxious and non-anxious participants at the AD stage (p = 0.0127).

**Figure 1.**
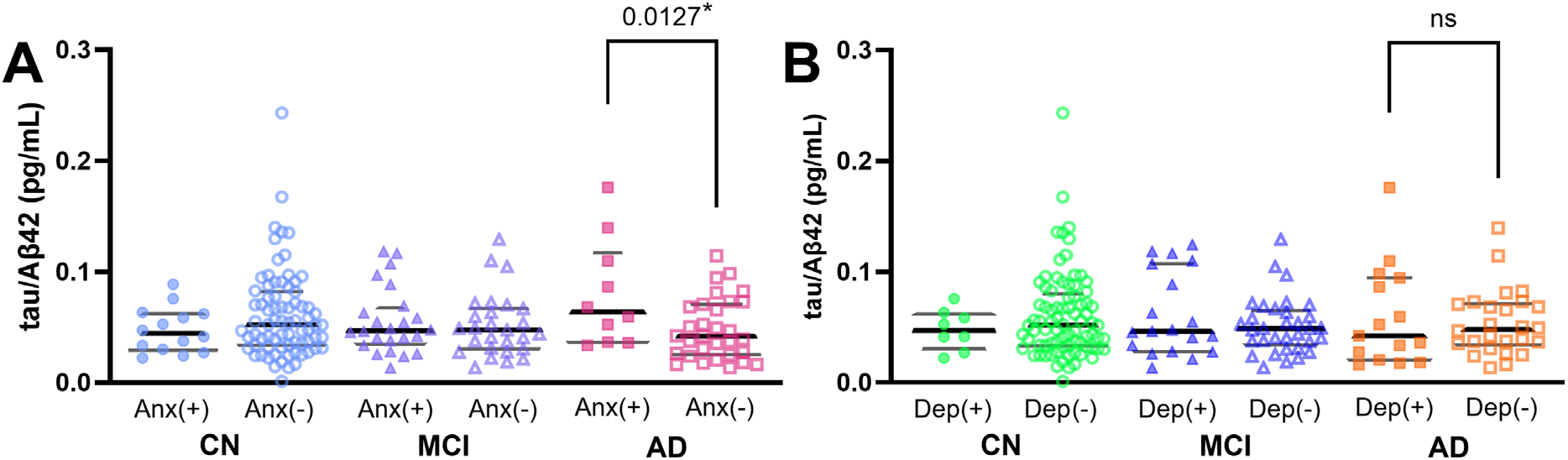
Comparisons of tau/Aβ42 levels in participants with(+) and without(-) A. Anxiety (Anx) or B. Depression (Dep) across stages. CN: Cognitively Normal, MCI: Mild Cognitive Impairment, AD: Alzheimer’s Dementia.

In comparison to anxiety, depression had a higher prevalence of 37% at the AD stage in our cohort. Despite this increased prevalence, however, we did not observe a significant difference in tau/Aβ42 ratio associated with depression at any stage (Figure 1B). Depression was also not associated with stage-specific differences in Aβ40, Aβ42, Aβ42/40, NfL, or tau (data not shown).

Lastly, we evaluated whether the higher tau/Aβ42 in the anxious AD group was associated with worsened cognitive performance (Table 2). Comparing psychometric performance on global (Mini-Mental State Examination) and domain-specific (animal and vegetable fluency, Trails A) tests, there was no significant difference between anxious and non-anxious AD groups.

## DISCUSSION

Our finding that plasma tau/Aβ42 was significantly elevated in anxious AD subjects is consistent with previous findings in CSF. In one longitudinal study of CN individuals, higher CSF tau/Aβ42 levels predicted a higher degree of anxiety and other NPS at 2-year follow-up [7]. In a prior cross-sectional study, it was observed that anxiety was significantly associated with lower levels of CSF Aβ42 and higher levels of t-tau, findings consistent with a larger tau/Aβ42 ratio [8]. Similarly to our findings, these results were not replicated in participants with depression.

Higher total tau levels are known to correlate with increased neurodegeneration [9], thus one interpretation of elevated tau/Aβ42 ratio is a greater degree of neurodegeneration at a given level of amyloidosis. This suggests that neurodegeneration takes place more quickly at a given disease stage for individuals with anxiety, and associates with a higher level of decline. Broadly, this is in agreement with previous literature. In one 3-year longitudinal study, among participants with MCI, the presence of anxiety led to an almost 2-fold increase in conversion to AD compared to non-anxious participants, and 30-fold in comparison to cognitively normal non-anxious participants [10]. Subsequent longitudinal studies have further confirmed these findings, with one study of 812 ADNI participants demonstrating NPI-Q reported anxiety at baseline significantly predicted MCI to AD conversion [11], and another study of 376 amnestic MCI participants observing significantly increased conversion to AD in anxious participants (with conversion increasing in line with anxiety severity) [12].

We have observed that this effect appears to be specific to anxiety, as our findings were not duplicated in participants with depression. This may suggest a pathophysiological mechanism or biomarker signature that is specific for anxiety. Alternatively, a recent review summarizes the evidence in the literature that depression in AD, in contrast to anxiety, may potentially arise more from psychosocial factors surrounding the disease, rather than a physiological mechanism [13]. Interestingly, we found that cognitive testing did not significantly differ between anxious and non-anxious AD participants, despite the significant difference in plasma tau/Aβ42. This finding adds further support to the theory that AD-related anxiety may not, in fact, be solely a psychosomatic response to the knowledge of cognitive decline, but instead perhaps has some heretofore undescribed physiological mechanism that is observable via biomarkers. Another possibility is that the rate of decline from the MCI stage to AD is increased in participants with anxiety [14], such that similar cognitive performance is reached over a shorter period of time.

Strengths of our study include a robustly phenotyped cohort, consensus diagnoses, and evaluation of plasma biomarkers via an ultrasensitive protein detection method. However, there are a few limitations. The cohort was modest in size and largely non-Hispanic White. The latter may lessen generalizability as NPS can have unique manifestations in different populations and associations with biomarker levels[15]. Further, our study evaluated NPS via the he NPI-Q, which is partner-based, and may differ from participant-based assessments.

In summary, this work demonstrates that anxiety is associated with unique plasma biomarker levels in AD, potentially related to increased neurodegeneration. This supports longitudinal investigation on the association of anxiety with biomarker changes and further study of the pathophysiologic relationship between anxiety and AD.

## ACKNOWLEDGMENTS

The authors have no acknowledgments to report

## DATA AVAILABILITY

The data supporting the findings of this study are available upon request from the corresponding author. The data are not publicly available due to privacy or ethical restrictions.

